# Quantitative Analysis of the Effectiveness of Public Health Measures on COVID-19 Transmission

**DOI:** 10.1101/2020.05.15.20102988

**Authors:** Thiago Christiano Silva, Leandro Anghinoni, Liang Zhao

**Affiliations:** Universidade de São Paulo, Ribeirão Preto, Brazil Universidade Católica de Brasília, Brasília, Brazil; Universidade de São Paulo, São Carlos, Brazil; Universidade de São Paulo, Ribeirão Preto, Brazil

**Keywords:** COVID-19, SARS-CoV-2, health policy, network, VAR, SIR

## Abstract

Although COVID-19 has spread almost all over the world, social isolation is still a controversial public health policy and governments of many countries still doubt its level of effectiveness. This situation can create deadlocks in places where there is a discrepancy among municipal, state and federal policies. The exponential increase of the number of infectious people and deaths in the last days shows that the COVID-19 epidemics is still at its early stage in Brazil and such political disarray can lead to very serious results. In this work, we study the COVID-19 epidemics in Brazilian cities using early-time approximations of the SIR model in networks. Different from other works, the underlying network is constructed by feeding real-world data on local COVID-19 cases reported by Brazilian cities to a regularized vector autoregressive model, which estimates directional COVID-19 transmission channels (links) of every pair of cities (vertices) using spectral network analysis. Our results reveal that social isolation and, especially, the use of masks can effectively reduce the transmission rate of COVID-19 in Brazil. We also build counterfactual scenarios to measure the human impact of these public health measures in terms of reducing the number of COVID-19 cases at the epidemics peak. We find that the efficiency of social isolation and of using of masks differs significantly across cities. For instance, we find that they would potentially decrease the COVID-19 epidemics peak in São Paulo (SP) and Brasília (DF) by 15% and 25%, respectively. We hope our study can support the design of further public health measures.

## 1. Introduction

The quick spread of the COVID-19 across countries has evidenced the high degree of interconnectedness worldwide. In less than six months, the COVID-19 epicenter traveled around the globe, starting in China, then moving to Italy, and to the US. The Coronavirus Resource Center at the John Hopkins University registers more than 4 million cases of the COVID-19 spread around 187 affected countries, i.e., roughly 96% of all countries recognized by the United Nations. Factors of such a rapid spreading include large flows of international air transportation, enabling cross-country jumps of the new coronavirus. Recently, the airline industry has been experiencing large drops in revenue mainly because of international border closures implemented by governments worldwide to detain “imported transmissions” of the virus. However, COVID-19 cases still substantially grow inside borders and represent a serious health concern of several countries across the globe. In this scenario, we can say that concerns about cross-country transmission have reduced and the understanding of the COVID-19 domestic transmission has gained much relevance.

This paper focuses on the COVID-19 domestic transmission in Brazil, which already registers cases in all 27 states as depicted in Figure 1a. We analyze the efficiency of public health measures—such as social isolation/quarantine and use of masks—in mitigating the COVID-19 transmission in the country using an innovative network-based approach that accounts for intra and intercity COVID-19 transmission channels. There are several unique features that make Brazil an important case study. First, there is a political confusion about the effectiveness of social isolation by the Brazilian federal and state governments [1]. The exponential increase in the number of infectious people and deaths in the last days indicates that such political disarray can lead to very serious results. Second, Brazil contains the 6th largest population in the world. Thus, the human impact of the COVID-19 can be substantial if not properly mitigated and a second wave of cross-country spillovers could be potentially sizable in the future.^1^ Third, Brazil has significant socioeconomic and cultural disparities across its 5,570 cities. Therefore, COVID-19 transmission and mortality rates may largely differ across cities, such as evidenced in Figures 1a–1b. The model proposed in this paper is able to estimate these city-specific COVID-19 transmission rates, thus accounting for their distinctive aspects. Fourth, WHO reports show that Latin America will most probably be the next epicenter of the COVID-19 outbreak. Since Brazil is the largest Latin American country and borders 83% of all South American countries, an understanding of the regional aspects of the COVID-19 transmission is crucial for designing public health measures.

**Figure 1:**
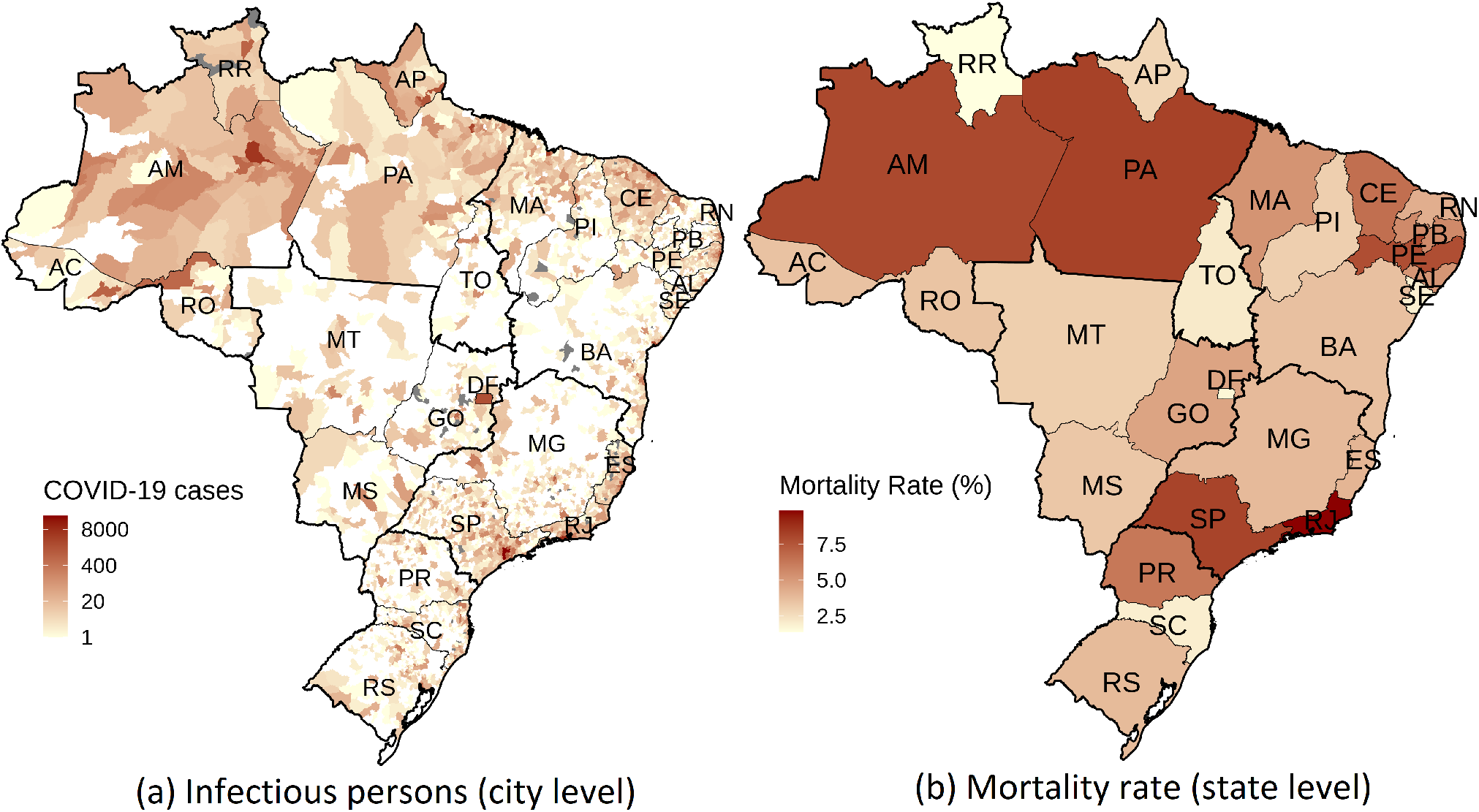
COVID-19 geographical spreading pattern in Brazil in terms of (a) COVID-19 cases at the city level and (b) mortality rate at the state level as of May 8, 2020. Gray areas represent cities that have not reported any COVID-19 epidemiologcal bulletin. We evaluate mortality rates by taking the ratio of the number of deaths due to COVID-19 to the number of infectious persons. Mortality rates are probably upward biased because the number of observed infectious persons is likely to be underestimated, as the COVID-19 may pass unnoticed for some cases (mild or no adverse conditions at all). We report mortality rates at the state rather than city level because there are many cities with few COVID-19 cases and deaths, which would distort the estimated mortality rates.

Most countries in the Americas are still facing the early stages of the COVID-19 and Brazil is no different. While it is important to have a full picture of the pandemic in each country to better design government policies aimed at mitigating the COVID-19 spread considering their local particularities, the omission of the government in taking effective measures at the onset of the epidemics can have large human and economic effects in the long term. Some eastern countries, such as China, South Korea and Singapore, may be an indication that having previous organized policies and mask usage culture are key to successfully mitigate the death toll. In this work, we consider only the availability of early-time data on the COVID-19 dynamics, thus better reflecting the real-world conditions that most governments are facing.

The dynamic of the COVID-19 epidemics is not only determined by the local aspects of cities. There is a continuous flow of persons from and to different cities either through roadways, domestic airlines, or sea routes that could transport the disease. However, these intercity transmission spillovers are not limited to biological risk factors. For instance, economic activities could also be related to the propensity of acquiring the virus from other places, such as when households or firms buy supplies abroad that are conditioned on surfaces that the virus is viable for long periods without proper sanitation.^2^

This transmission dynamic renders each city subject not only to its inherent “COVID-19 natural transmission rate” dictated by the local aspects of the city itself—such as demography, culture, law, and weather—but also from outside the city. Our model permits to estimate transmission rates of each city while accounting for infectious factors from the outside using the Susceptible-Infectious-Recovered (SIR) model in a special type of transmission network among cities.

We take an innovative approach to construct the underlying COVID-19 transmission network among cities. Even though we apply the model for the COVID-19 propagation inside Brazil, the model is general and could be applied for any networked environment, such as in cross-country studies or even more granular approaches than at the city level. We model such network using a weighted directed graph. Vertices are cities and links represent potential COVID-19 contagion/spillovers from one city to another. To estimate the links, we consider a panel-format data^3^ composed of city-specific COVID-19 infectious counts of locals over time. We then use a vector autoregressive (VAR) model to find directional COVID-19 transmissions of every pair of cities in the network. Since the seminal paper of [5], VARs have provided key empirical input into substantive economic and financial aspects. Despite the robustness of the model, their use in epidemiology is still a new topic. Here, we design a VAR model that explicitly considers the temporal ordering of the disease spreading. We let every city-specific infectious count be dependent not only on its own past value but also from all other cities. The weights of past values of each city *j* that influence the current city *i*’s infectious local count are the links in our network. Such links are estimated by fitting the entire network structure to temporal city-specific infectious count data. We mitigate concerns with parameter overfitting by using an elastic net regularization scheme during training time^4^ and one-step ahead rolling validation methodologies borrowed from the machine learning literature.^5^

An interesting property of the early-time dynamics of a SIR model is that it still enables us to estimate the transmission rate *β* of the model. Given the recovery rate *γ* of infectious persons,^6^ then the model can be completely described [11], including late-time dynamics and infectious peak. It is worth mentioning that the rate *γ* can be divided into two parts, the time from onset to death and the time from onset to recovery. Both can vary from country to country, since they are highly correlated to demographics, health care system and the treatments available. The onset to recovery time is, however, invariant to the topological structure of the system and, therefore, we use an average value of 14 days in all scenarios of our study

In early-time dynamics, the effective transmission rate *β* of an *isolated SIR* and a *networked SIR* model differs by the spectrum of the estimated COVID-19 transmission network. When we do not consider the network environment, we are effectively supposing the existence of a single large city composed of all cities in the model. In this way, the susceptibility of being infected depends on the total number of infected (all cities). The introduction of multiple cities effectively reduces this propensity by imposing that the likelihood of being infected is higher inside cities rather than across cities. The network spectrum corresponds to the largest eigenvalue of the network adjacency matrix. If the isolated SIR has a transmission rate *β*, then the networked SIR will have an effective transmission rate of *β*_eff_ = *λ*_max_*β*, in which *λ*_max_ is the largest eigenvalue of the network. The network spectrum encodes all the graph structure in terms of its ability of spreading and amplifying intercity contagion at early time.

In this paper, we also analyze the efficiency of health policy measures implemented by the Brazilian government to mitigate the COVID-19 propagation. Social isolation and quarantine measures were adopted by several states at different time scales. Following that, the Brazilian Health Ministry recommended the use of masks at the federal level. Political disagreements on the effectiveness of quarantine measures by the federal and state governments were on display and may have lead the population into confusion, thus affecting the efficacy of such measures. Our work contributes to this discussion by estimating the joint efficacy of these measures.

We find that the quarantine and use of masks measures decreased the growth rate of the spectrum of the COVID-19 transmission network over time, suggesting that the measures were effective. To get a sense, Figure 2 portrays the average COVID-19 growth rate of cities in the state of São Paulo segregated in terms of their average social distancing index in the period.^7^ First, after the use of masks recommendation, the COVID-19 growth rate, in general, decreased. However, it decreased more in cities of São Paulo with low social distancing measures. This may be due to the fact that these cities could have more potential close human-to-human contact and therefore the use of masks is crucial to detain the COVID-19 transmission. To get a sense of the human impact of such measures, we build counterfactual scenarios in which we consider that none of these measures were taken by the government. By running the SIR model in networks, we find that the quarantine and the use of masks recommendation reduced the peak of the COVID-19 epidemics, on average, in 15% in São Paulo (SP) and almost 25% in Brasília (DF), when we look at the average effect in the last week of available data (May 2 to 8, 2020). This reduction is explained by the flattening of the epidemics curve: São Paulo (SP) and Brasília (DF) have peak date shifts from July 7 to July 24 and August 29 to September 28, respectively.

**Figure 2:**
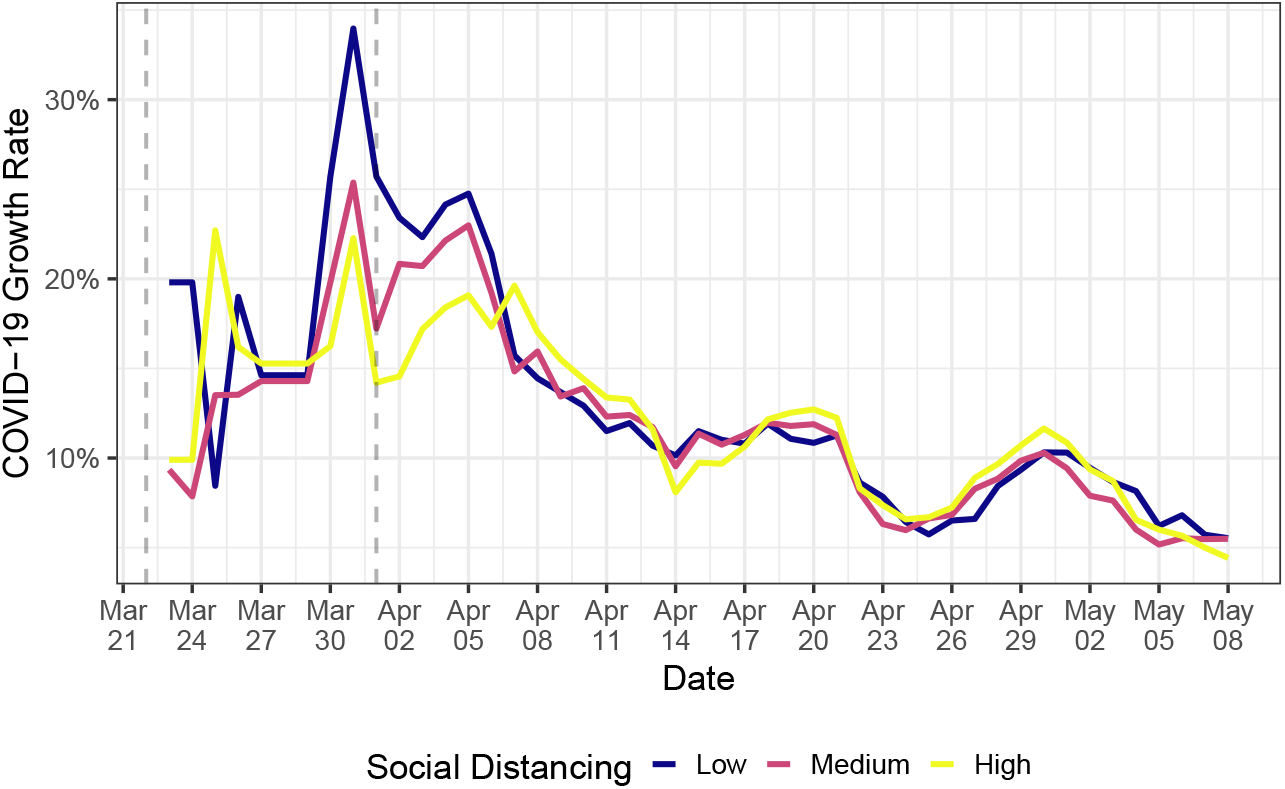
Average COVID-19 growth rate in cities of the state of São Paulo, Brazil, with low, medium, and high social-distancing indices. Available data goes until May 8, 2020. The first vertical line is the beginning of SP quarantine, while the second represents the use of mask recommendation by the federal government. Data from social distancing is public and comes from the São Paulo State Government (in Portuguese). To alleviate week seasonality, we use 7-day moving averages to construct the average growth rates. The low, medium, and high social-distancing indices represent the bottom, middle, and upper terciles of the corresponding distribution. Data from the number of infectious persons per each city is discussed in Section 4.1.

Our results show the increasing trend of infectious cases in the last days, which is confirmed by the updated official data in Brazil. This situation is consistent with the decreasing social isolation rate shown by Figure 2, which, in turn, probably caused by the political discrepancy in public health measure application.

## 2. Related background and literature

In this section, we present relevant background on SIR models in networks and the related literature about our work.

### 2.1. Relevant background: early-time dynamic of SIR models in networks

In this section, we present relevant background on the Susceptible-Infectious-Recovered (SIR) model in networks. We refer the reader to [11] for a comprehensive analysis on epidemiological models and to [12] for the seminal paper on the original SIR model. Since we focus on the early-time dynamics of the SIR models, we can assume that the number of births and deaths are much smaller than the population, in a way that the closed population hypothesis holds.

Define as ***s**_i_*(*t*), ***x**_i_*(*t*), and ***r**_i_*(*t*) the share of susceptible, infectious, and recovery persons of city *i* relative to the local population at time *t*. In a closed population, the SIR model in networks is government by the following differential equations:

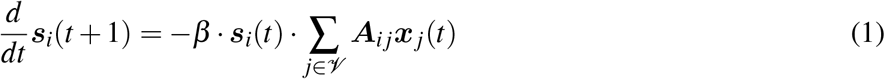

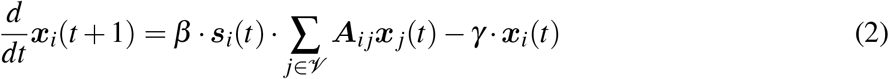

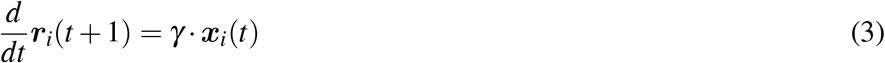

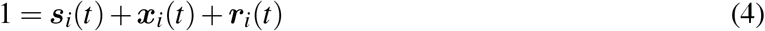

∀*i* ∈ *V* and *t* ≥ 0. We can substitute (4) into (2), yielding:

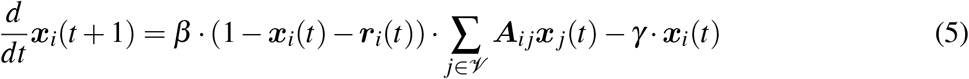

In early time, i.e., we can assume that ***x**_i_*(*t*) ≪ 1 and ***r**_i_*(*t*) ≈ 0, *∀i ∈ 𝓋*. Therefore, we can ignore second-order ***x**_i_*(*t*) terms and effectively set ***r**_i_*(*t*) to 0. With these modifications, Equation (5) becomes:

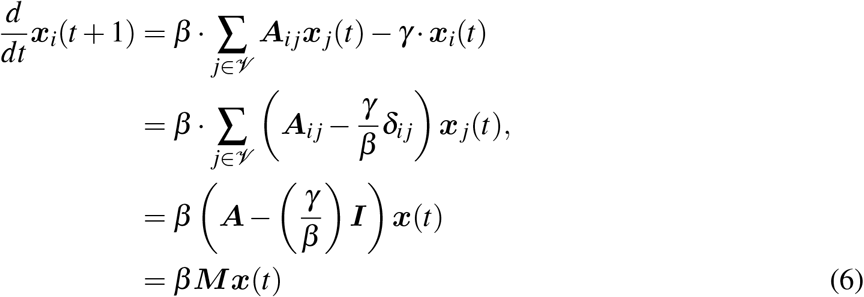

in which ***I*** is the identity matrix, 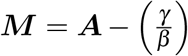 is the adjacency matrix ***A*** with a homogeneous perturbation of 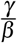 in the main diagonal, and *δ_i__j_* = 1 if *i* = *j*, and *δ_i__j_* = 0 otherwise. Equation (6) is a standard differential linear system whose solution can be written in terms of the eigenvector basis of the adjacency matrix ***A***:

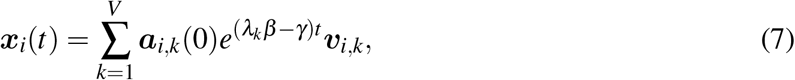

in which ***A · v**_k_* = *λ_k_**v**_k_* holds *k* 1*, …,V*. The term *λ_k_* is the *k*-th eigenvalue of ***A**, **v**_i,k_* is the *i*-th entry of the eigenvector associated with the *k*-th eigenvalue. The parameter ***a**_i,k_*(0) in (7) is a scaling constant that depends on the initial condition of city *i*.

In early time, the growth rate of equation (7) is government by the exponent term with the largest eigen-value *λ*_1_ = *λ*_max_ of matrix ***A***, which is a well-known measure from spectral graph theory denominated graph spectrum [13]. Therefore:

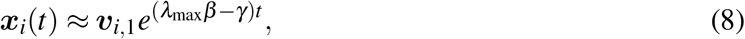

− i.e., the growth rate is *λ*_max_*β* − *γ* and the probability of contagion is proportional to the eigenvector associate with the largest eigenvalue *λ*_max_, ***v***_1_, which corresponds to the eigenvector centrality measure of the graph, according to the spectral graph theory [13].

### 2.2. Relative literature

Basically, there are two strategies to prevent epidemic spreading in networks [14]. One is the efficient immunization protocols and the other is to find out relevant spreaders and activation mechanisms.

Immunization strategies are methods for identification of nodes that shall be immunized, taking into account the network structure. Immunized nodes and all the incident links can be removed from the epidemic network. Immunization can not only protect immunized individuals, but can also reduce the epidemic threshold, precluding the outbreak of the disease. Among various immunization strategies, random immunization protocol is the simplest one, where a fraction of randomly selected nodes are made immune. However, in this case, the immunization threshold tends to be 1 in heterogeneous networks, indicating that almost the whole network must be immunized to suppress the disease [15]. Target immunization protocol considers special nodes to be immunized. In [16, 15], the authors show that the immunization threshold can be exponentially small over a large range of the spreading rate if considers the immunization of a fraction of nodes with the largest degree. Other approaches consider not only the critical nodes, but also the entire prevalence curve (the so-called viral conductance) [17, 18].

Although immunization is a fundamental strategy in the epidemic study, the research community pays also much attention to find out which nodes, links and local structures are most influential or effective in the spreading process [19, 20, 21, 22, 23, 24, 25, 26]. These findings aimed at understanding network measures on nodes and links, such as degree, betweenness, K-core index, closeness, link property on spreading dynamics. Besides of finding superspreaders, some researchers also worked on the identification of how topological features influence global epidemics [27, 28, 29].

However, the above mentioned strategies require the discovery of vaccine or at least partial knowledge on the epidemic network under consideration. With the mass and quick spreading of COVID-19, neither of them is a practical method to prevent the outbreak. Therefore, global intervention methods, like social isolation, even lockdown, have already been proven to be efficient. For this reason, we study the effectiveness of public intervention methods. Our results provide strong evidence on the effectiveness of public health measures, such as quarantine and use of masks, to reduce the increasing rate of infection even without detailed information of the highly dynamical population network.

## 3. Methodology

This section discusses the underpinnings of our methodology. Our analysis consists of the following stages:

1. *Network construction*: we construct the COVID-19 network transmission network by fitting the network links to real data.
2. *COVID-19 epidemics estimation using the SIR model*: we use the network estimated in Step 1 and simulate the COVID-19 evolution in every city of the network.
3. *Effectiveness evaluation of public health policy*: we change the network structure so as to simulate the omission of public health policies and run our epidemics model in Step 2 without the government intervention. We estimate the efficiency of the public health policies by inspecting the change in the COVID-19 epidemics peak.

### 3.1. Network construction using panel data

Consider the weighted directed graph *ℊ* = *𝓋, ℯ* in which *𝓋* is the set of vertices and *ℰ* is the set of links. There are *V* = *𝓋* vertices and *E* = *ℰ* links in the network. In our epidemiological application, vertices can represent cities, states, countries, or any well-defined entity or geographical circumscription (neighborhood, street, house etc.). For simplicity and with no loss of generality, we denominate the vertices as cities. We assume as given the set of cities/vertices *𝓋*. In contrast, links between cities *i* and *j* connote potential COVID-19 transmission from *i* to *j* and are *a priori* unknown. In the context of cities, city-to-city contagion could happen for a series of reasons, such as when infectious persons visit or migrate or even from intercity transportation of supplies covered in surfaces that the SARS-CoV-2 is viable for long periods. Therefore, the network *𝒢* encodes all potential transmission paths between cities be through organic or non-organic media. The goal of this section is to estimate the set of links *ℰ*, i.e., the intercity COVID-19 transmission channels.

Let ***x***(*t*) = [***x***_1_(*t*)*, **x***_2_(*t*)*, …, **x**_V_* (*t*)] denote the vector with shares of infectious persons relative to the local population of every city *i ∈ 𝒱* in the network at discrete time *t* ≥ 0. Specifically, we denote as ***x**_i_*(*t*) ∈ [0, 1] the share of infectious persons within city *i* at time *t*. That is, we take the ratio between the number of infectious persons to the total local population in the city. When ***x**_i_*(*t*) = 1, then all population in the city is infectious. When ***x**_i_*(*t*) = 0, none is infectious. In-between values represent partial shares of infectious population. Define the column vector ***x**_i_* = [***x**_i_*(0)*, **x**_i_*(1)*,…, **x**_i_*(*T*)]^′^ as the COVID-19 time series evolution in city *i* up to time *T*, in which the superscript^′^ is the transpose operator. Since we perform an early-time analysis of the epidemics, *T* is likely to not be large. Let also the matrix ***X*** = [***x***_1_*, **x***_2_*, …, **x**_V_*], dim(***X***) = *T × V*, be all the cities’ time series with the shares of infectious persons stacked in columns over all period with available data (panel data).

To construct the network, we consider the temporal ordering of the COVID-19 spread across different cities. We attempt to describe the current share of infectious persons vector ***x**_t_* with the same vector immediately at the previous time step, i.e., ***x***_*t*−1_ as follows:

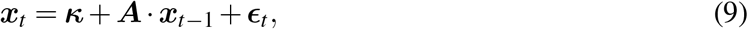

∀*t* ∈ {0, 1*, …, T*. The term ***κ***, dim(***κ***) = *V* × 1, is an intercept column vector; ***A***, dim(***A***) = *V ×V*, is the adjacency matrix encoding the set of links *ℰ* of the graph; and ***∊**_t_* (**0, Σ_*∊*_**) is the unobservable zero mean white noise vector process (serially uncorrelated or independent) with time-invariant covariance ∈ matrix **Σ_*∊*_**. Let ***A**_i__j_* be the (*i, j*)-entry of ***A**, i, j ∈ 𝒱*. When ***A**_i__j_ >* 0, then city *i* can spillover COVID-19 to city *j*. The larger ***A**_i__j_* is, the stronger is such contagion. Then, the set of links is given by *ℰ* = {*i, j* ∈ *𝒱* : ***A**_i__j_ >* 0}.

The terms ***κ**, **A*** in Equation (9) are unknown and are estimated using a fitting process to the observed data ***X***. Equation (9) describes a VAR(1) model. To ensure that the system is stable, the companion matrix must have roots inside the complex unit circle. To guarantee such property, our variables ***x**_i_, i* ∈ *𝒱*, must be stationary. Since they are lower- and upper-bounded—i.e., ***x**_i_* ∈ [0, 1]—then they are stationary by construction. Specifically, we minimize the following regularized loss function *L* [7] using the coordinate descent algorithm [30]:

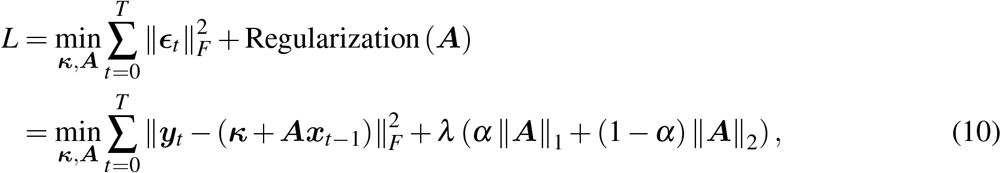

in which *λ* ≥ 0 is the *elastic net* regularization term and *α* ∈ [0, 1] is the tradeoff parameter between Lasso (*L*_1_ norm) and Ridge (*L*_2_ norm) regularizations. We notate *∥·∥*_*F*_, ∥·∥_1_, ∥·∥_2_ as the Frobenius, *L*_1_, and *L*_2_ norms, respectively. Larger values of *λ* encourage sparser networks. The first term represents minimization of the error term ***∊**_t_, ∀t* ∈ 0, 1*, …, T*, and ensures that the estimated adjacency matrix ***A*** better reflects the COVID-19 transmission dynamics over time. The second term is a regularization term over the adjacency matrix ***A*** introduced to prevent overfitting and ensure that the estimation is numerically tractable. We do not regularize the intercept vector ***κ*** because it conceptually adapts to the city-specific average values of our data.

There is an empirical challenge in fitting the adjacency matrix ***A*** to the panel data ***X*** when we are dealing with large-scale networks in which the number of cities *V* largely surpasses the number of available time points *T*, i.e., when *V ≫ T*. Such problem is aggravated when we only have early-time information about the disease, i.e., *T* is small. In this case, we would incur in overparametrization and overfitting is a concern. The regularization term in (10) mitigates such concern. We opt for an *elastic net* regularization scheme because it is a robust regularizator that combines positive features of Lasso and Ridge regularizations [30].

Due to the temporal dependency of the panel data, the usual *k*-fold cross-validation is not well-suited for our model selection procedure. Following [7], we optimize the penalty parameters *λ* and *α* in (10) using a *h*-step ahead mean-square forecast error (MSFE). Due to data availability, we keep *h* = 1 so as to minimize further data losses. We divide the data into three equally-spaced and contiguous periods: (i) initialization (*t* ∈ {0*,…, T*_1_}), (ii) training (*t* ∈, {*T*_1_ + 1*, …, T*_2_}), and (iii) forecast evaluation (*t* ∈ {*T*_2_ + 1*, …, T*}), in which 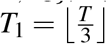 and 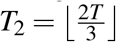. We also use a rolling validation process as follows. We first fit the model using all data up to time *T*_1_ and forecast 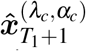, in which *λ_c_* and *α_c_* are fixed candidate penalty terms. We then sequentially add one observation at a time and repeat this process until *T*_2_ − 1. Then, we choose the penalty terms *λ* and *α* that minimize the one-step ahead MSFE given by:

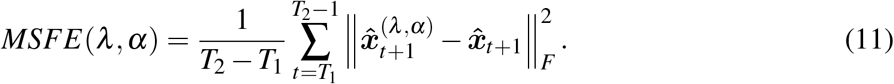

Finally, we estimate the one-step ahead forecast accuracy using data points in *t* ∈ {*T*_2_*, …, T*}, which have not been used in the model selection procedure. To better assess the potentiality of the network in amplifying contagion across different municipalities, we remove the self-loops in the estimated network, which correspond to the influence of the local infectious population on its own future value.

### 3.2. Estimating transmission rate in early-time epidemics networks

In this section, we assume the network structure *𝒢* = *𝒱, ℰ* as given, i.e., the set of vertices and links are already established in accordance with the network construction described in Section 3.1. We start from the results of the early-time dynamic of SIR models in networks described in Section 2.1. Therein, we show that the growth rate at early time is determined by *λ*_max_*β − γ* (see Equation (8)). Therefore, the graph spectrum *λ*_max_ modulates the transmission rate parameter by either amplifying or dampening the contagion speed.

If *λ*_max_*β > γ*, then Equation (8) grows exponentially, while it decays when *λ*_max_*β < γ*. Therefore, the reproduction number (critical point) is 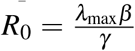. Recall that the reproduction number in the SIR model without network is 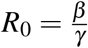[12]. Therefore, the reproduction numbers of both models differ by the graph spectrum of ***A**, λ*_max_.

Equation (8) assumes that every city in the model has a single growth rate dynamics dictated by the term *λ*_max_*β − γ*. Changes in the epidemics spreading for each city would then be fully determined by their eigenvector centralities, because growth rates are identical across cities (see Equation (8)). However, studies show that the transmission rate parameter *β* is dependent on local aspects of cities [11]. In contrast, the recovery rate parameter is much less variable across different places. As mentioned earlier in this study, WHO indicates an average recovery time of 14 days for mild cases. Therefore, we consider a different transmission rate for each city in the network *β_i_* while letting fixed the recovery rate *γ* for all cities. We can still apply the classical framework of SIR in networks because, even though transmission rates are city specific, they tend to be normally distributed around some mean natural value. That is, large deviations are unusual. We empirically find this fact using our application to the Brazilian case. Mathematically, we rewrite (8) as follows:

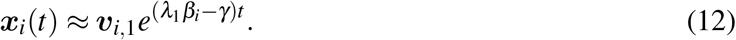

We can linearize (12) by simply taking the log(.) at both sides of the equation for each city *i* in the network:

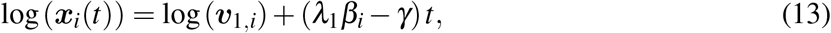

*∀i ∈ 𝒱*. The LHS and RHS are always non-negative, because ***x***(0) ≥ 0 and is non-decreasing (early-time assumption), *e*^(*λ*^1*βi*−*γ*)*t* ≥ 0 (asymptotically speaking), and ***v***_1*,i*_ ≥ 0 [13]. We can then apply the log(.) without any restrictions. We can estimate (13) for all cities *i* at once by adding dummies for the constant and time-dependent term for each city in the model (2 dummies per each city). We end up with a set of 2*n* – 1 dummy variables, because the last one is the reference dummy. Since we have a panel data with temporal dependencies (the same city appears multiple times), we use a linear panel-data estimation model [31] as follows:

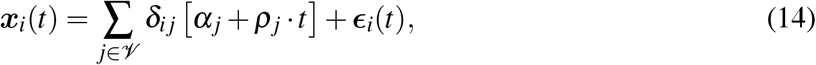

*∀i ∈ 𝒱*, in which *α_i_* and *ρ_i_* are the constant and time-variant dummy terms for city *i*, and *ε_i_*(*t*) is the residual from the least square estimation with dummies. We cluster the errors at the city level, such as to mitigate concerns with heteroskedasticity and serial correlation, which could bias our coefficient estimates. Equations (13) and (14) are linked by the following identities:

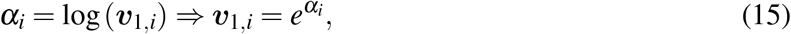

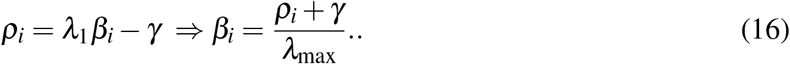

Given the recovery rate *γ*—which is assumed to not change over time nor across cities—we can fully identify the eigenvector centrality and the local transmission rate of every city *i* using (15) and (16), respectively. We only take city-specific estimations of ***v***_1*,i*_ and *β_i_* that are statistically significant at the 10% level. Otherwise, we set the estimated coefficients to zero.

### 3.3. Assessing efficiency of health policy measures in epidemics spreading

With our framework, we can analyze the speed of the epidemics spreading through the network at early time by simply inspecting the graph spectrum *λ*_max_ = *λ*_1_ for different time horizons using the methodology described in Section 3.1. Since the reproduction number of the epidemics is proportional to the graph spectrum, then large graph spectra indicate a higher speed of contagion. Any changes of the graph spectrum can be attributed to a “net effect” of public policies of the government in the entire network. Since we use the share of infectious persons of each Brazilian city, then this “net effect” comprises not only federal policies, but also state- and even city-level policies.

Moreover, we can estimate the human impact of these policies in terms of changes in the number of infectious persons at the peak by running the SIR model described in Section 3.2 for each estimated city-specific transmission rate parameter *β_i_* defined in (16) and for different values of the graph spectrum. We use a conservative approach and compare the largest observed graph spectrum with the most recent graph spectrum in our dataset. We assume that the largest graph spectrum occurs when public policies were still latent and were not having effects in the epidemics spreading. Most recent values of the graph spectrum are assumed to represent transmission dynamic after public policies were in, as was the case in Brazil who adopted quarantine and recommended the use of masks in the period that we have available data.

## 4. Application

In this section, we apply our model to Brazilian data at the city level.

### 4.1. Data

We use daily data on the number of infectious persons per each city in Brazil using COVID-19 epidemiological bulletins of 27 State Health Departments from February 25, 2020, to May 8, 2020.^8^ Each Brazilian state compiles local reports from cities inside their geographical circumscription. We end up with 60,021 city-time epidemiological bulletins comprising 2,754 (out of 5,570) cities affected by COVID-19 in Brazil.

Our data is representative because local hospitals are required by law to register any COVID-19 events to the local government while cities and states must notify the federal government. However, there may be substantial sub-notifications due to persons that acquire the COVID-19 and recover unnoticed or without hospitalization.

We also collect city-level population estimates in the Brazilian Institute of Geography and Statistics (IBGE), which is the agency responsible for official collection of statistical, geographic, cartographic, geodetic and environmental information in Brazil. We evaluate the share of infectious persons by taking the ratio of COVID-19 cases reported in the local health bulletin and the local population size. The use of shares in our estimation models is important because it is a stationary variable.

We apply a three-day smoothing filter on the number of infectious persons in each municipality to alleviate concerns with late contamination reports or short-term rectifications by the local health government that could compromise our estimations. In our network construction procedure (see Section 3.1), we keep only cities that reported COVID-19 cases in at least 20% of the available time frame. Our results remain qualitatively the same if we do not apply this filtering criterion. In our estimation of the SIR parameters (see Section 3.2), we center all time points in relation to the occurrence of the first death in the city.

Figures 3a–3b portray the COVID-19 evolution in six of the most affected cities in Brazil relatively to the first reported death in terms of the number of COVID-19 cases and as a share of the local population size, respectively. São Paulo (SP) has the highest number of infectious persons. However, there is strong size effect: São Paulo (SP) has almost 12.2 million residents while the second largest city, Rio de Janeiro (RJ), has almost half of that (6.8 million). To get a sense of the local COVID-19 criticality, we can look at its evolution as a share of the local population. In this case, we note that COVID-19 transmission speed is much larger in Manaus (AM) and Fortaleza (CE). Brasília (DF) and Porto Alegre (RS) have smaller transmission rates and local COVID-19 criticality. However, mortality rates may not follow such incidence criticality, because they correlate with local health quality and demography characteristics.

**Figure 3:**
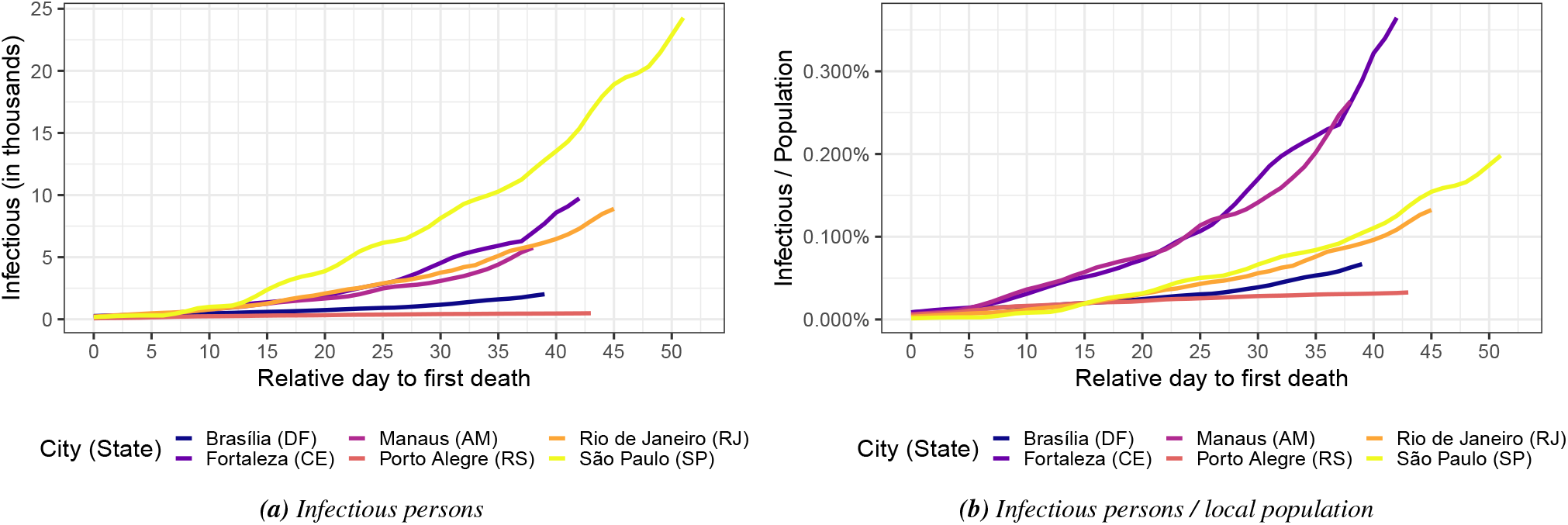
COVID-19 evolution in six of the most affected cities in Brazil (a) in absolute terms (number of infectious persons) and (a) as a share of the local population size. Horizontal axis represent the relative day in terms of the first observed death due to the COVID-19.

### 4.2. Results

This section presents the main empirical results of the paper. We first build the COVID-19 intercity transmission network and analyze its propensity of amplifying the COVID-19 in different cities. Then, we analyze the net effectiveness of public health measures adopted by the Brazilian government.

#### 4.2.1. Intercity COVID-19 transmission network in Brazil

Figure 4 shows the graph spectrum of the COVID-19 intercity transmission network of Brazil over time. For each time point (horizontal axis), we run the network construction through the fitting process in Section 3.1 with data from the beginning of the sample up to that specific time point. Even though our sample starts in February 25, 2020, we start the fitting process from March 13, 2020, such as to have enough data for the fitting process. That is, we start with 18 time points for each Brazilian city. Therefore, we initially divide the panel data in three equally-sized groups with 6 time points for model training, model selection (parameters and penalty terms), and model evaluation. These group sizes increase as we add more time points. We perform the network construction estimation daily from March 13 to May 8, 2020, in an independent manner.

**Figure 4:**
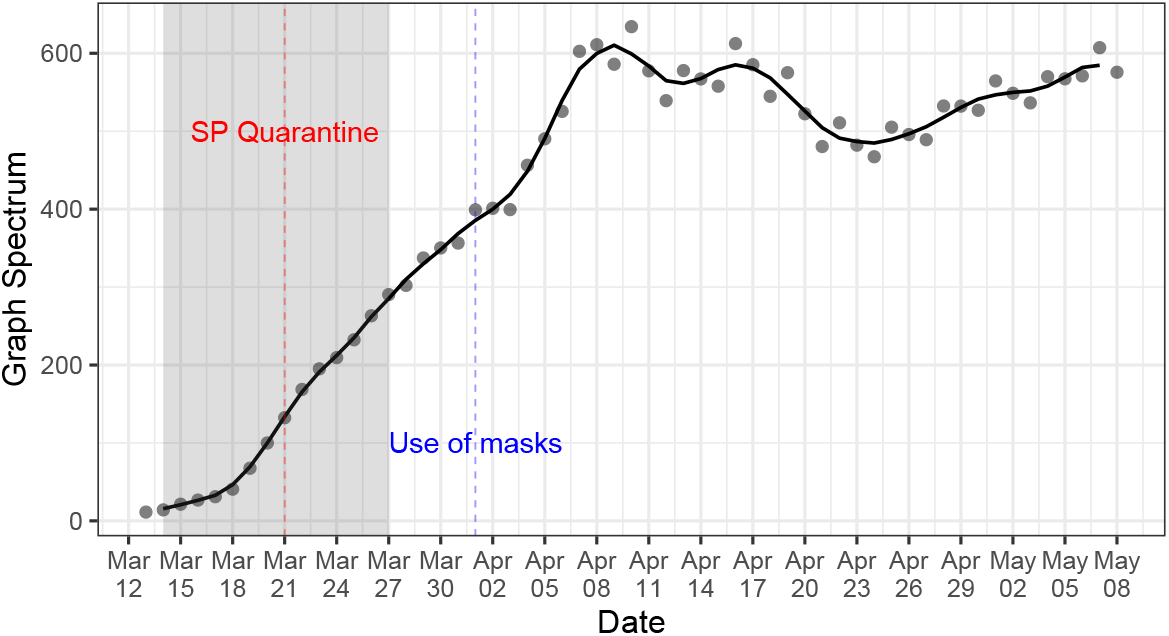
Graph spectrum of the COVID-19 intercity transmission network of Brazil (see Section 3.1). The shaded area indicating the timing window in which quarantine measures were adopted by the most affected Brazilian states. The red dashed line indices the beginning of the quarantine in São Paulo, the COVID-19 epicenter in Brazil. The blue dashed line indices the beginning of the use of masks recommendation by the Federal Health Ministry. For each time point (horizontal axis), we build the network with city-specific shares of infectious persons with data up to that point.

In Figure 4, we add a shaded area indicating the timing window in which quarantine measures were adopted by the most affected Brazilian states. Since São Paulo is the COVID-19 epicenter in Brazil as it encloses 57.4% of all the COVID-19 infections in Brazil, we also add a vertical dashed red line indicating the beginning of the quarantine adopted by the São Paulo State Government. We also draw the use of masks recommendation beginning date by the Federal Health Ministry in Brazil as a dashed blue line enacted. While quarantine measures are at the state level, the use of masks recommendations goes at the federal level and encompasses all the 5,570 cities and 27 states in Brazil. São Paulo is the most central city in the transmission network. Therefore, it practically shapes the graph spectrum of the intercity transmission network.

We observe a reduction in the growth rate of the graph spectrum after the quarantine measures precisely two days after the measure. However, the growth rate still persisted at positive rates, indicating that the COVID-19 transmission speed kept increasing after such measure, but with a slower pace. After the incubation period following the use of masks recommendation, we observe a drastic change in the graph spectrum. The growth rate changed sign and started to reduce, showing that the set of health policy measures taken by the government was efficient. However, after April 23, 2020, the graph spectrum again started to increase. This can be due to several factors, such as social confusion in following health guidelines in view of the political disarray that Brazil is facing, or even non-compliance with quarantine and use of masks measures. Our model does not permit to have an isolated causal impact of the use of masks recommendation nor of the quarantine measures. However, it enables us to understand how the set of all policy measures affected the COVID-19 transmission rate across cities over time. Combining Figures 2 and 4, it seems that the reduction in the COVID-19 growth rates after the use of masks recommendation was more apparent in cities with relative low social distancing indices. This may be due to the fact that these cities have more potential close human-to-human contact and therefore the use of masks is crucial to detain the COVID-19 transmission.

To understand the topological aspects of the COVID-19 intercity transmission network, Figure 5 plots the PageRank centrality for the top 5 most central cities in each of the five regions in Brazil. We normalize the PageRank with respect to the most central city: São Paulo (SP) on May 8, 2020. As the city centrality becomes higher, the more it contributes to spreading the COVID-19 throughout the network. The top 5 most central cities in the country are the following state capitals (in decreasing order): (i) São Paulo (SP), (ii) Rio de Janeiro (RJ), (iii) Fortaleza (CE), (iv) Recife (PE), and (v) Manaus (AM). These cities all have airports and are strongly interconnected to the remainder of cities in Brazil through roadways and are likely to be the hubs for the COVID-19 spread to other nearby cities in Brazil, especially countryside municipalities. The centrality of São Paulo (SP) in the Southeast monotonically increases over the entire sample. The same roughly occurs with Manaus (AM) in the North, Fortaleza (CE) in the Northeast. Porto Alegre (RS) in the South and Brasília (DF) in the Midwest have the highest centralities in their region but with a negative growth rate in the last days of the sample. Overall, there is a very heterogeneous profile of the city centralities over time, showing the underlying non-trivial patterns in the COVID-19 transmission network.

**Figure 5:**
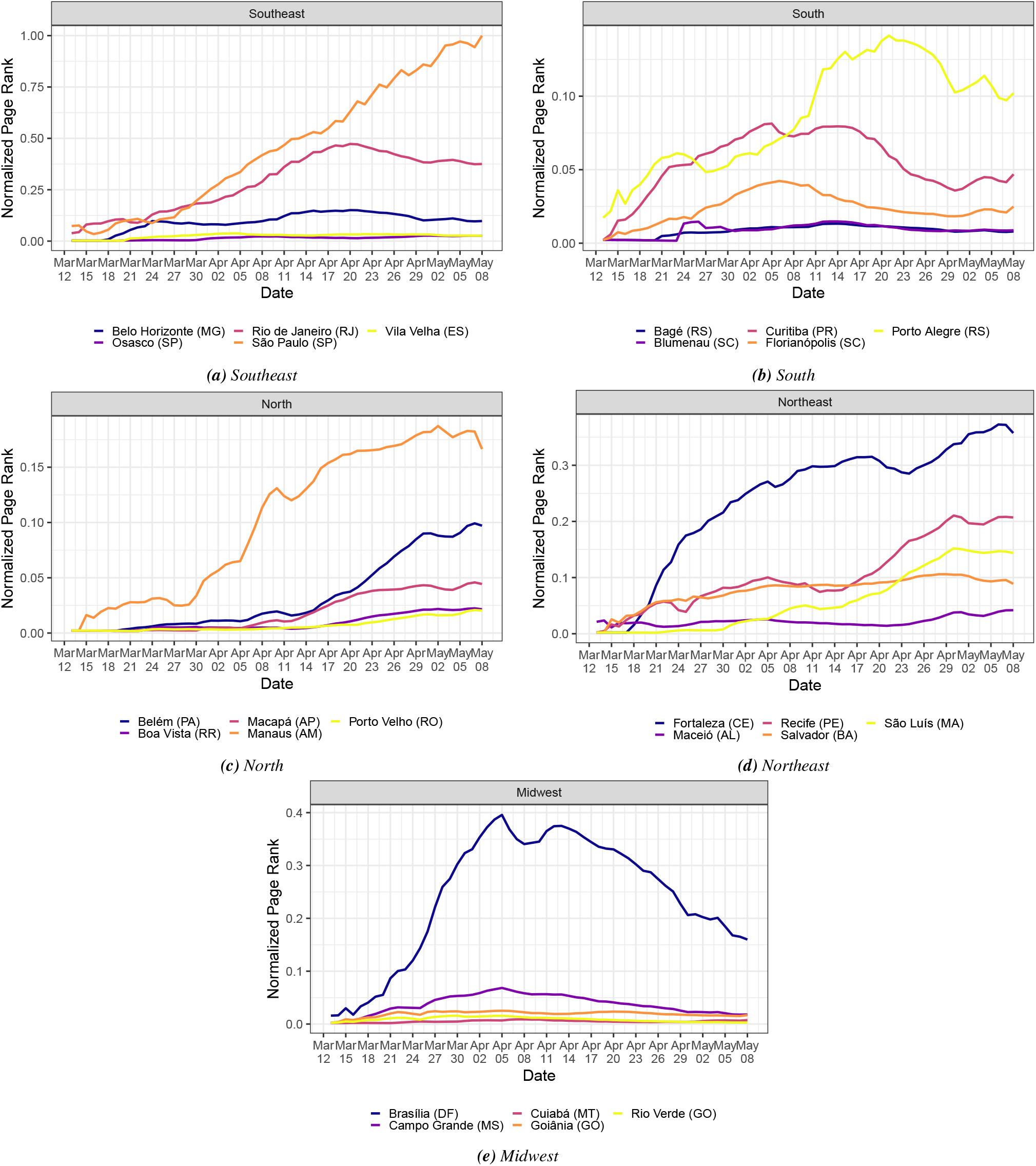
Evolution of the normalized PageRank centrality measure in the COVID-19 transmission network (see Section 3.1 for the network construction details). We only report the top 5 cities with highest PageRank at each Brazilian region. For each time point (horizontal axis), we build the network with city-specific shares of infectious persons with temporal data up to that point. Each label is composed of the city name followed by its state inside parentheses.

#### 4.2.2. Measuring the human impact of health policy measures to mitigate the COVID-19 propagation

In this section, we run the SIR in networks (see Equations (1)–(4)) with different transmission rate parameters for each city in Brazil, in accordance with (16). We first estimate the city-specific *ρ_i_* using the panel-data information on counts of the share of infectious persons in each city in Brazil via (14).

Then, we estimate the transmission rate parameter *β_i_* of each city *i ∈ 𝒱* in Brazil by fixing the recovery rate parameter as *γ* = 1*/*14. We use the remaining parameter *λ*_max_—the graph spectrum—to evaluate the effectiveness of the set of health policy measures in detaining the COVID-19 in Brazil. We take as baseline model the graph spectrum reached in April 10, 2020, which is the maximum observed value. We assume that this graph spectrum would have not changed afterwards in case the set of health policy measures were not taken.^9^ We then run several SIR models with the observed graph spectrum values in Figure 4 after the graph spectrum maximum in April, 10, 2020.

Figure 6a shows a comparison of the infectious peaks of the baseline SIR model—i.e., the hypothetical scenario in which health policy measures were not introduced—and the ones with graph spectrum values observed daily after that maximum. The vertical axis shows the relative change in these infectious peaks of the baseline and the observed model day by day, which can be interpreted in terms of the potential share of spared infections at the infectious peak due to the introduction of the set of health policy measures. Since we have data from each city affected by the COVID-19, we plot the median, percentiles 75% (0.25 distant from the median) and 90% (0.40) of this distribution. In the Supplementary Material, we provide the effectiveness of public health policies for each affected municipality in Brazil. In April 10, 2020, the share of spared infections in the epidemics peak is zero, because the baseline model is compared with itself. Then, as we move forward in time and use smaller graph spectrum values, as shown in Figure 4, the potential share of spared infectious increases. The share of spared infectious persons in the epidemics peak reaches a median value 40% lower than that of the baseline model when we use the graph spectrum in April, 24, 2020, suggesting high effectiveness of the quarantine and use of masks health policies. After this point, the share of spared persons decreases— reflecting the increase in the graph spectrum in Figure 4—giving more room for the spread of the COVID-19. The effectiveness of the health policy measures, however, remains positive throughout the entire sample.

**Figure 6:**
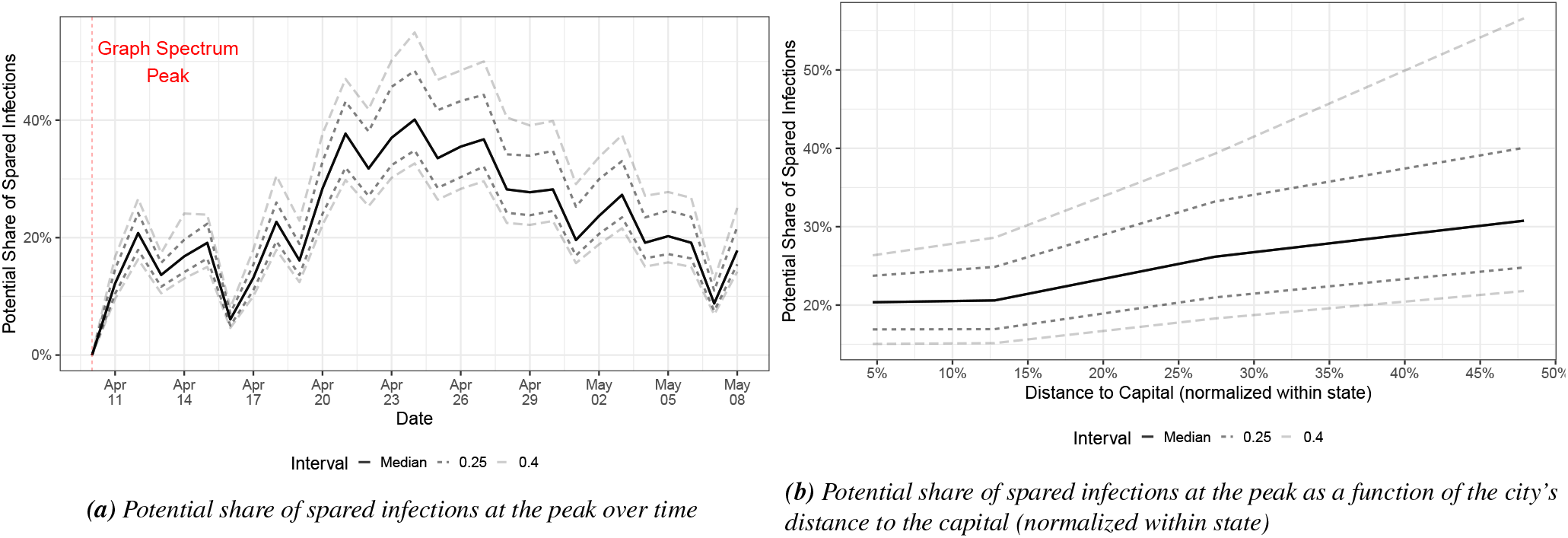
Distribution of the efficiency of health policy measures along all affected cities in Brazil over time. We plot the efficiency distribution as a function of (a) time and (b) the city’s distance to the capital within the same state it resides. Since states in Brazil have substantial differences in their sizes, we normalize the city’s distance to the capital to the most distant city within the state.

The first case of the COVID-19 in Brazil was reported in São Paulo (SP) on February 25, 2020. After that, it spread to several Brazilian state capitals probably through air transportation (most of the airports in Brazil are in the state capitals and capitals are far from each other). The epidemics took some time before reaching the first case in countryside cities. Figure 6b displays the distribution of the potential share of spared infections in terms of the city distance to the state capital. Since Brazilian states are very different in size, we normalize the distance to the most distant city within the state. We observe a positive relationship between potential share of spared infectious and distance to the capital, suggesting that health public policies are most effective in cities that are distant from the capital. This may reflect not only the temporal delay of the COVID-19 in reaching the countryside, which puts the local COVID-19 at very early time in these regions, but also demography aspects, such as lower population density, and agricultural economic activities that do not require large conglomerates of persons.

Figure 7a shows the effectiveness of the set of public health measures for six of the most affected Brazilian capitals. In particular, Brasília (DF) reaches a 50% lower share of infectious persons at the peak when we compare peaks reached with the graph spectrum value on April 24, 2020 (against the baseline in April 10, 2020). Figure 7b shows the number of potential spared infectious persons due to the set of health policy measures. This figure is constructed by simply multiplying the share of spared infectious with the local population size of each of the six cities. Since São Paulo (SP) is the largest city, it would potentially spare more persons when the COVID-19 epidemics reach its peak.

**Figure 7:**
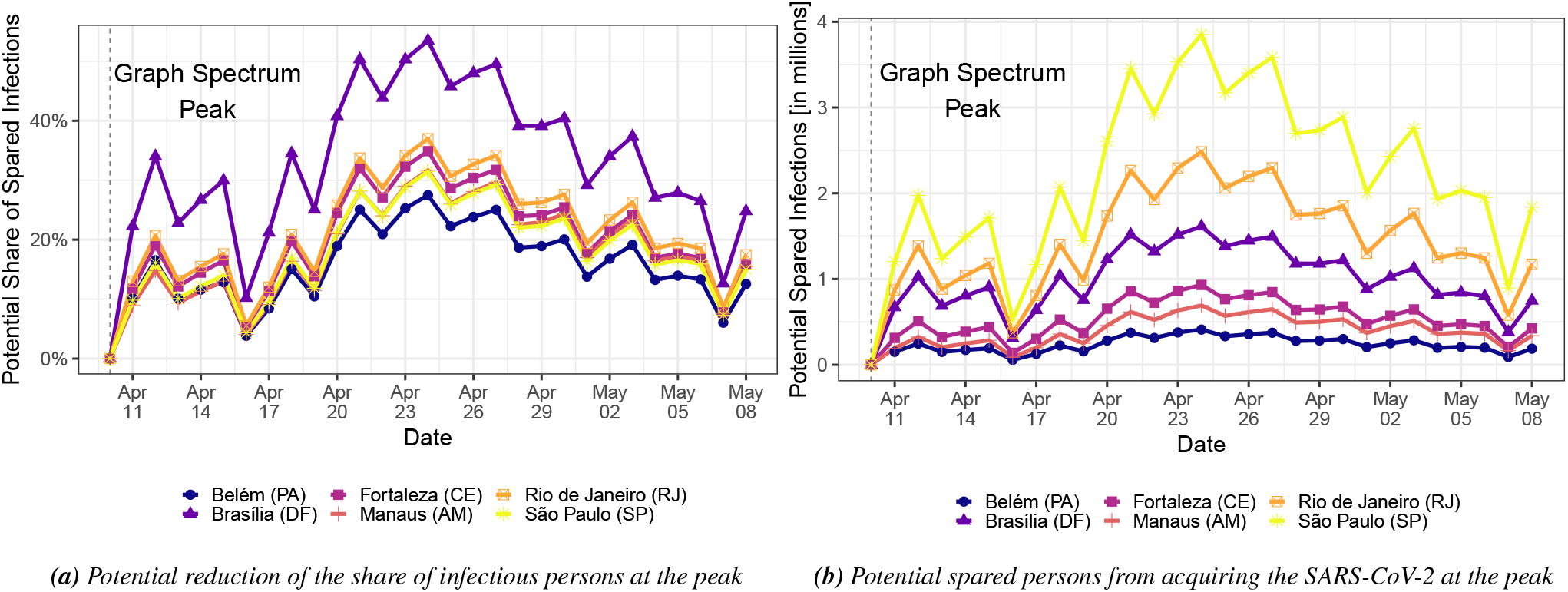
Efficiency of public health measures over time as a function of (a) the share of the spared local population and (b) the spared number of persons (in millions). We depict curves only for six capital cities that are being substantially affected by the COVID-19: Beleém (PA), Fortaleza (CE), Rio de Janeiro (RJ), Brasília (DF), Manaus (AM), and São Paulo (SP).

## 5. Conclusions

At the current stage of the COVID-19 infection, many countries have stopped the entrance of foreigners. Therefore, the study of virus transmission dynamics inside each country gains relevance. In the last few days, Brazil has become one of the most infectious countries in the world. In this work, we present a general epidemics transmission model and apply it to the Brazilian case. Our method has three steps. First, we construct the COVID-19 transmission network by fitting city-specific COVID-19 cases over time to calibrate the network links, which represent intercity COVID-19 transmission. Second, we gauge the network propensity of spreading COVID-19 throughout the cities using a spectral graph analysis. Third, we propose a methodology to quantify the effectiveness of public health policies using the dynamics of early-time SIR model and spectral network theory.

Our spectral network analysis indicates that social isolation and the use of masks can effectively reduce the transmission rate of the COVID-19 in Brazil. The COVID-19 propagation dynamics seems to decrease following these public health policies when we also consider an incubation period, which lags the effect of any COVID-19 mitigation measure. Moreover, our empirical analysis supports the view that use of masks seems to be more effective than social isolation, which is further corroborated by what is being occurring in Austria [32]. With no vaccine up to date, public health intervention is still the main method of epidemic control. We hope our study can help the government make correct decisions.

## Data Availability

All data used in this work is public available.

## Acknowledgements

This work is supported in part by the São Paulo State Research Foundation (FAPESP) under grant numbers 2015/50122-0 and 2013/07375-0, the Brazilian Coordination of Superior Level Staff Improvement (CAPES), the Pro-Rectory of Research (PRP) of University of São Paulo under grant number 2018.1.1702.59.8, and the Brazilian National Council for Scientific and Technological Development (CNPq) under grant numbers 303199/2019-9, 308171/2019-5, and 408546/2018-2.

## Authors contributions statement

T.C.S., L.A. and L.Z. conceived the study idea and researched the related work. T.C.S. designed the method and conducted the experiments. T.C.S. and L.A. gathered the datasets. L.Z. supervised the research scheme. All authors reviewed the manuscript.

## Supplementary Material

This Supplementary Material presents additional results of our empirical application to Brazil.

**Table A1:**
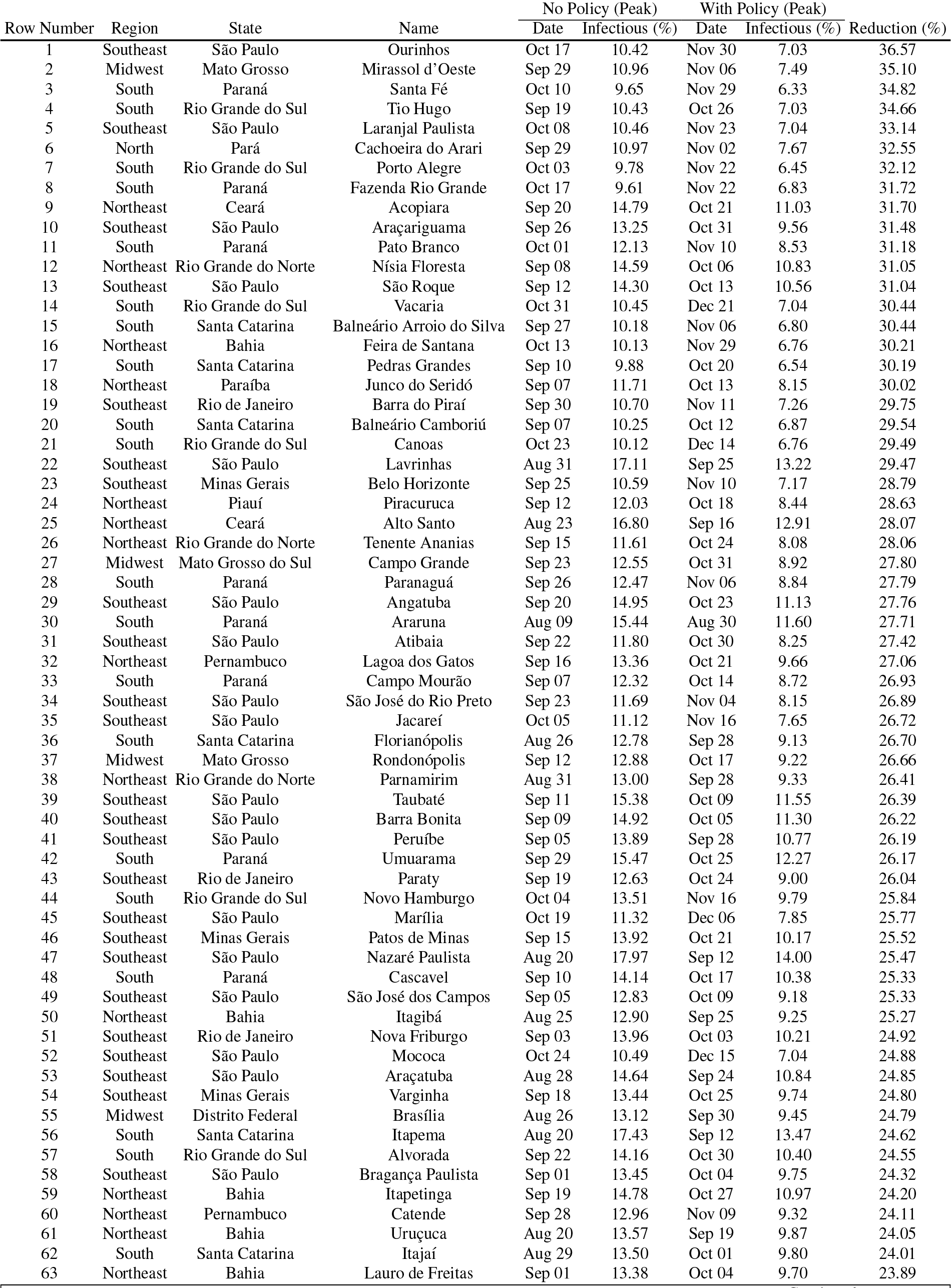
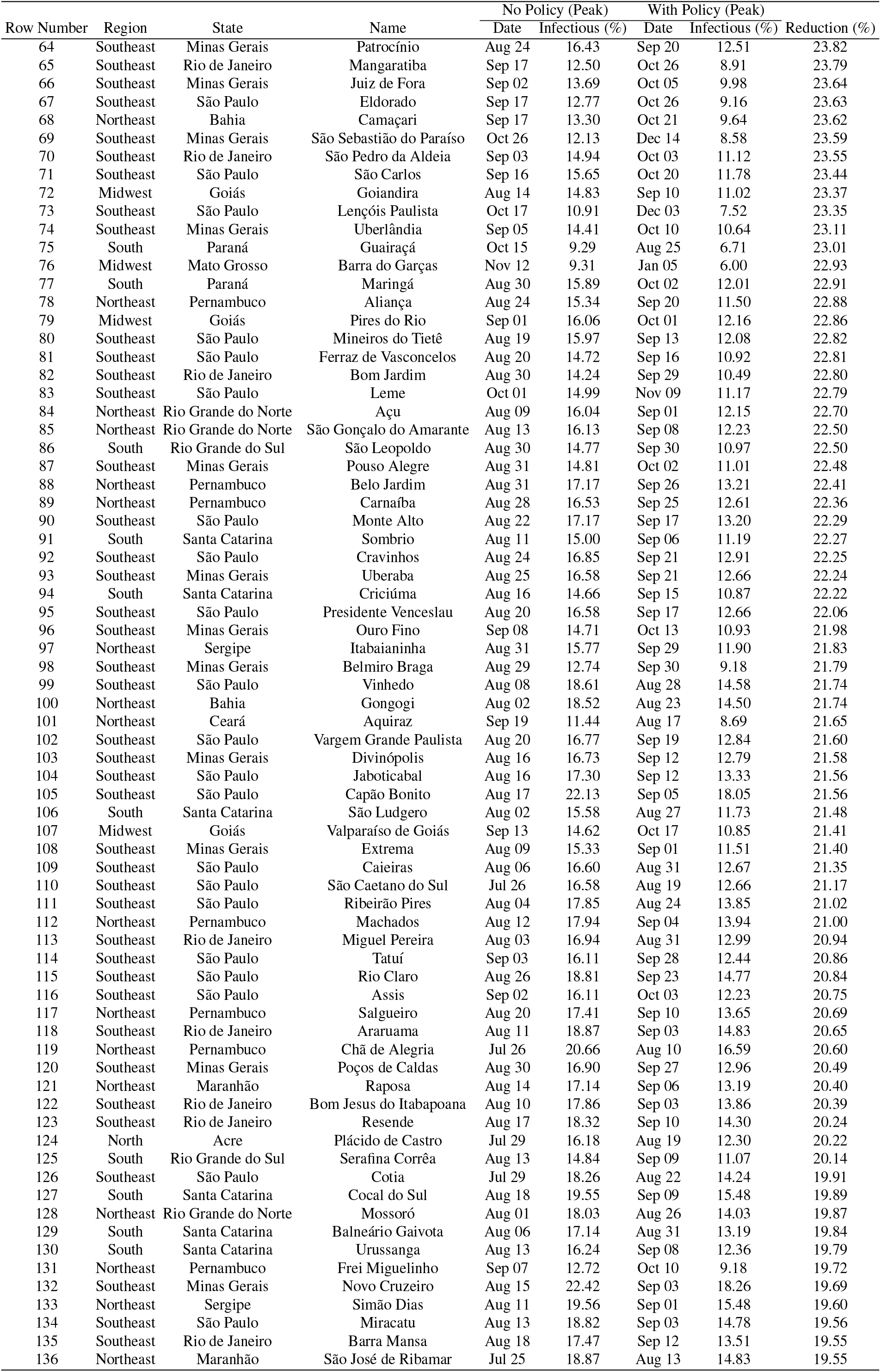
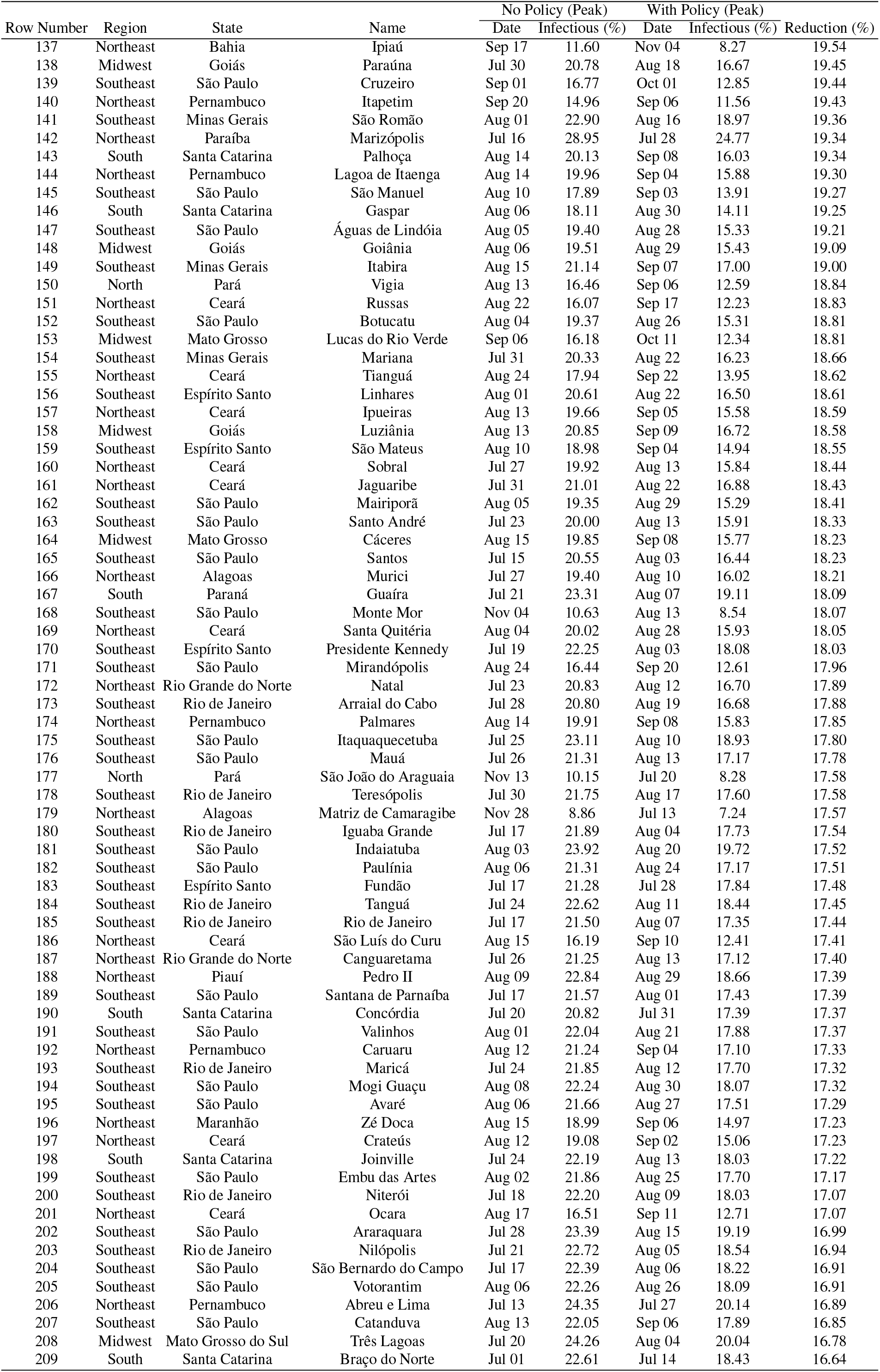
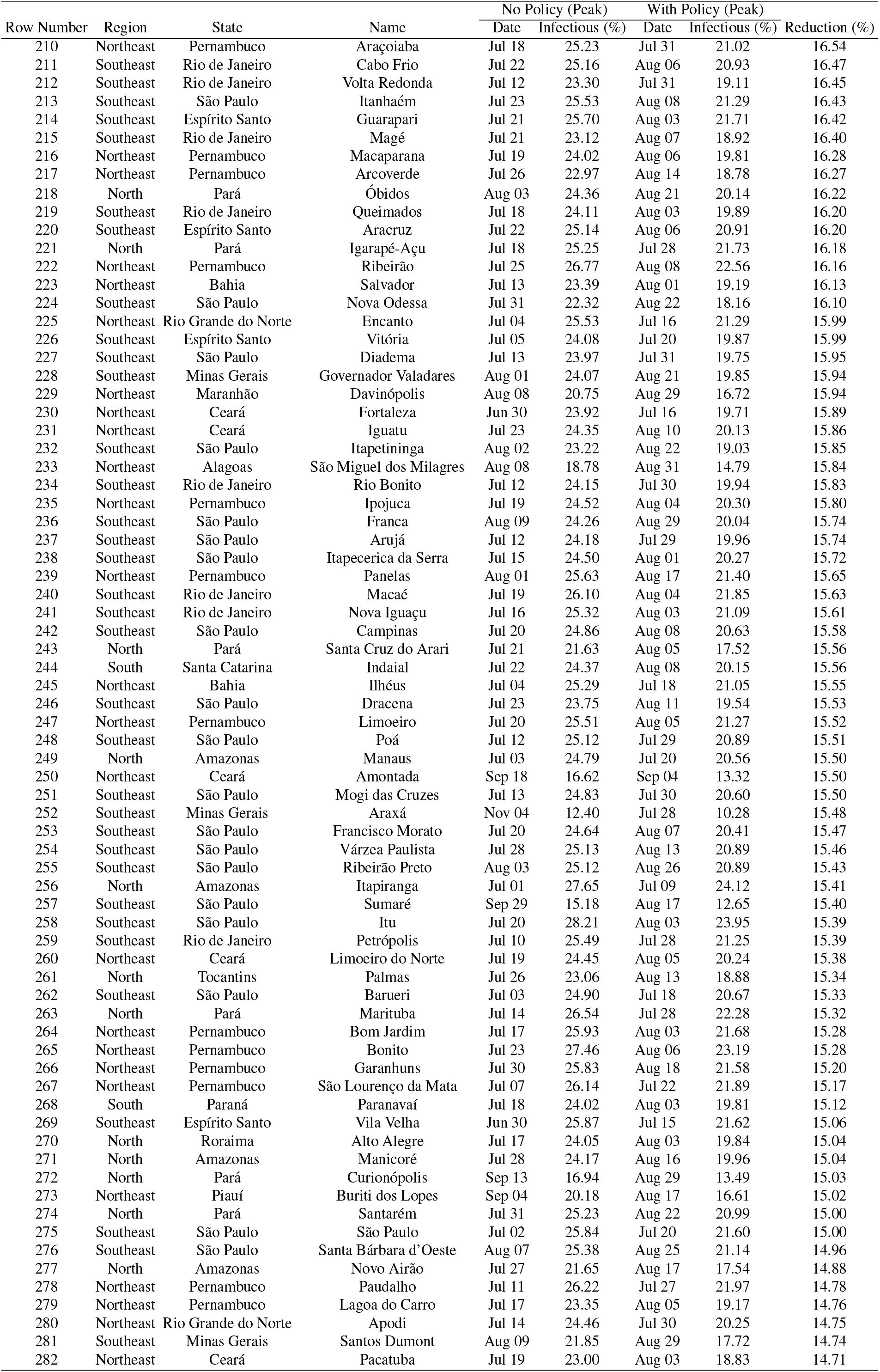
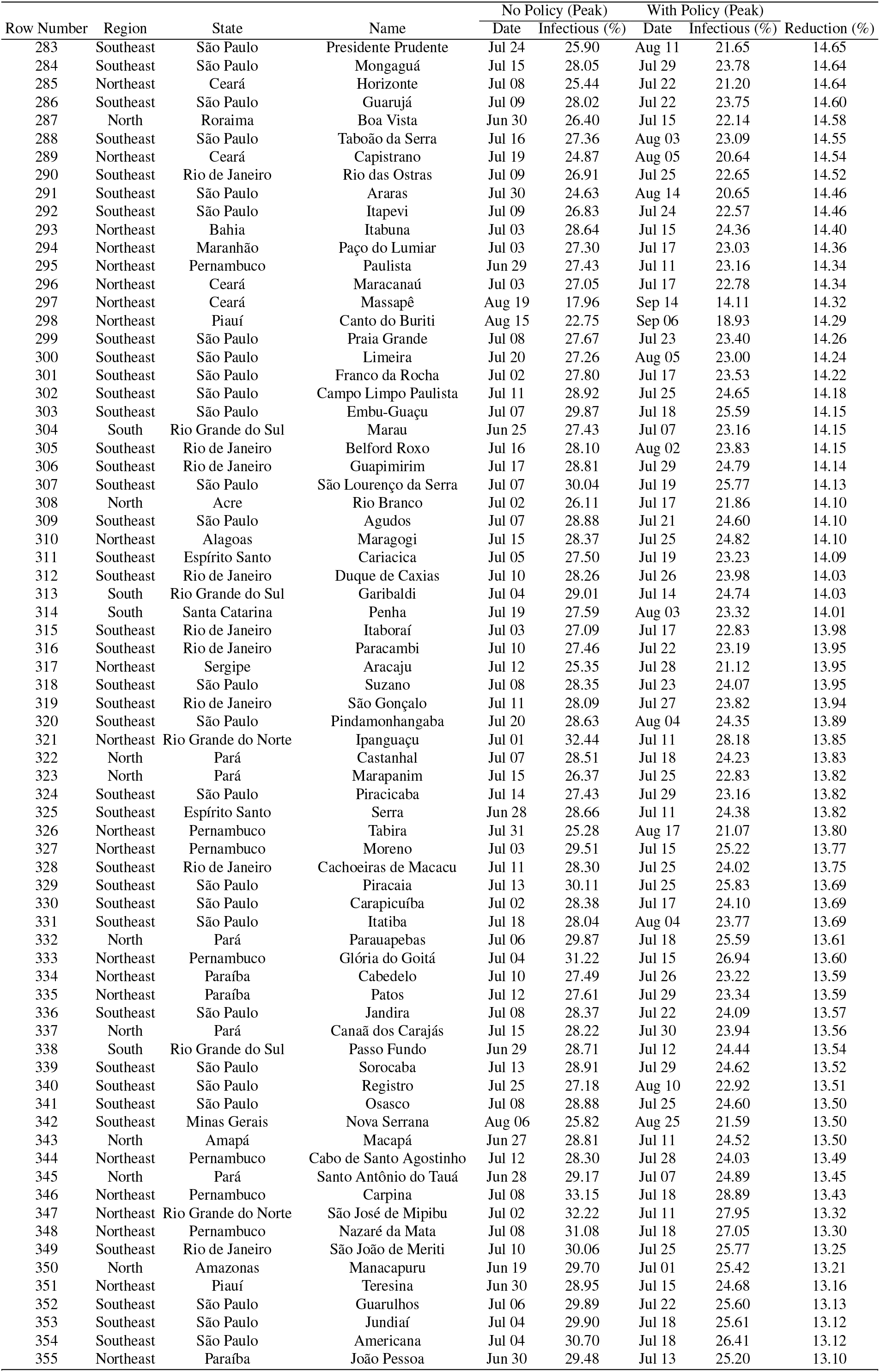
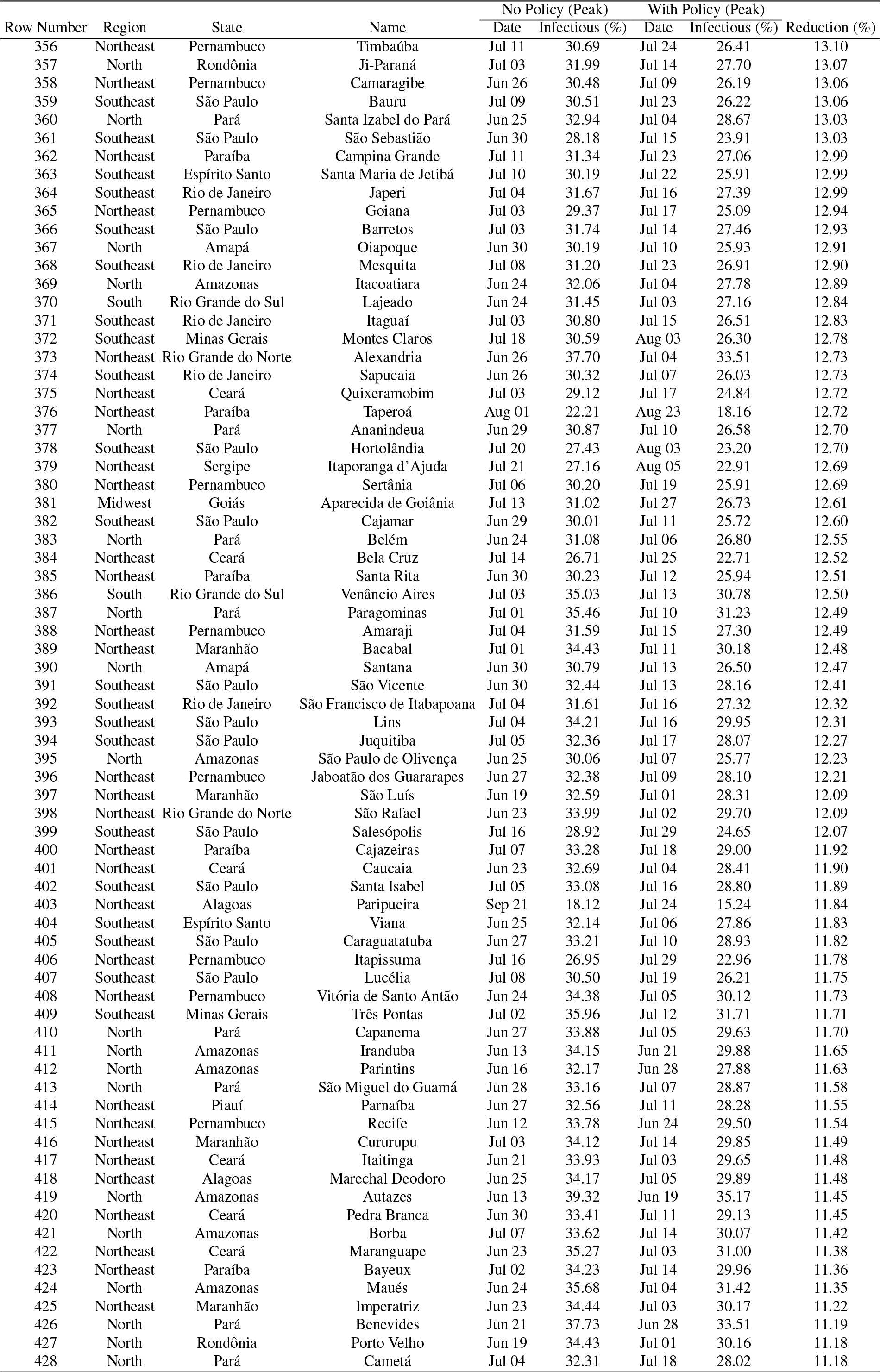
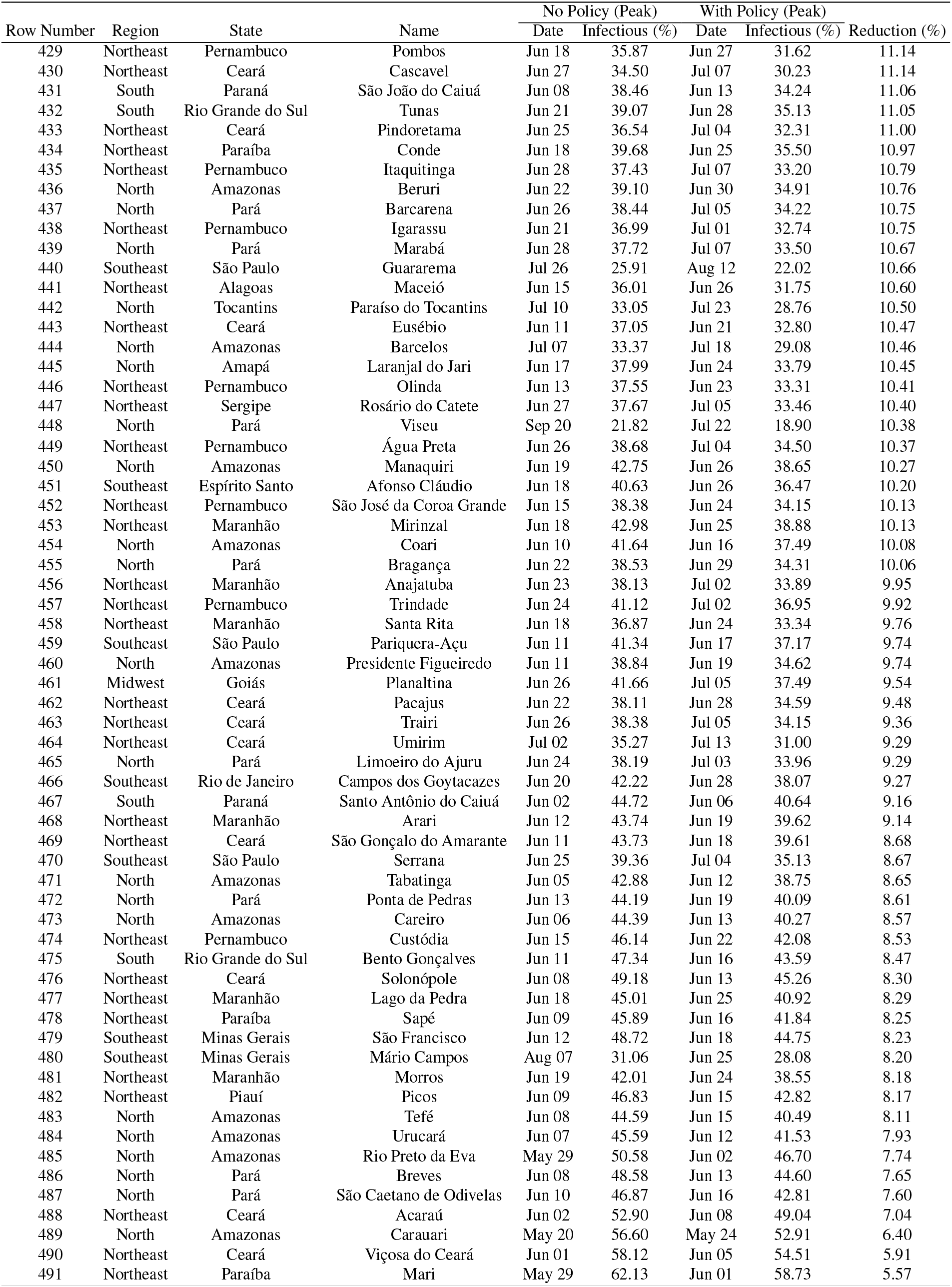
Estimated share of the local population with COVID-19 in Brazil at the peak and the corresponding month and day in 2020. We report the peak date and share of infectious persons to the local population of the city with and without health policy measures (see Section 4.2 for details). This simulation uses data up to May 8, 2020. We only report estimates for cities in which the simulated infectious peak with policy is higher than 5% of the local population.

## Footnotes

1 the authors projects recurrent wintertime outbreaks of SARS-CoV-2 will occur after the initial pandemic wave. They argue that prolonged or intermittent social distancing could be necessary up to 2022. Even with apparent elimination, the authors state that the resurgence in contagion could be possible as late as 2024.

2 Studies have show that the virus is more stable on smooth surfaces, such as plastic and stainless steel (detectable up to 7 days), and is very sensitive to temperature (the inactivation time is reduced to 5 mins at 70 degrees Celsius)[3]. The aerosol and surface stability of SARS-CoV-2 is similar to SARS-CoV-1, with a half-life of about 1hr in the form of aerosol and up to 7hrs on plastic surfaces. Other surfaces, such as copper, cardboard and stainless steel have also long half-life values, ranging from 1 to 6 hours [4].

3 A *panel data* is composed of *n* multivariate time series, each representing the evolution of COVID-19 cases of a specific city. It is a mixture of *cross-sectional data*—in which we observe *n* cities all in a specific time point—and *time series data*—in which we observe a single individual over time.

4 The elastic net is composed of a convex combination of the Lasso (*L*_1_) and Ridge (*L*_2_ norm) regularization. We refer the reader to the seminal work of [6] for further details.

5 Parameter overfitting becomes a serious concern when we have several cities in the model. For instance, we apply our method to Brazilian data, which is a country with vast territorial dimensions and with more 5, 570 cities (end of 2019). In this case, we would have to estimate 5, 570 5, 570 31 million parameters with only a few time points (because we only have early-time data). Ensuring regularization is vital to have reasonable out-of-sample estimates. See [7] for more details on regularization of VAR models.

6 The recovery rate can be estimated from the timeline between the appearance of symptoms and the case resolution. Several ongoing studies report estimates for the recovery rate. For instance, the authors in [8] assumes that the duration of the infection ranges from 15 to 20 days. Data from the outbreak in Wuhan show an onset-to-death time of 17.8 days and an onset-to-recovery time of 24.7 days [9]. This results are, however, biased to higher values due to the overwhelmed health care system in Wuhan in the early days of the outbreak and the sub-notification of the outcome of mild-cases. Reports from WHO indicate a recovery time of 14 days for mild cases and 21–42 days for severe cases. Among those who die the onset to outcome ranges from 14 to 56 days [10]. Since the mild cases account for most of the cases, we set gamma to 14 days in this study.

7 Such index represents the extent of compliance of the population to the quarantine measures.

8 This data is scattered around a large quantity of state government sites. In general, the bulletins are not standardized across different states and not even cities. We use the compiled dataset from Brasil.io for this task.

8 Asymptomatic and mild-cases can represent up to 80% of the cases according to China reported numbers. This cases tend not to be tested in Brazil.

9 This is a conservative approach, because we can observe a positive momentum of the graph spectrum growth rate prior to reaching April 10, 2020. However, we cannot be sure whether such graph spectrum would still increase if these policies were not in place. Therefore, we keep the conservative approach and consider such point as the maximum.

